# Anti-tetanus vaccination is associated with reduced occurrence and slower progression of Parkinson’s disease

**DOI:** 10.1101/2024.05.03.24306800

**Authors:** Ariel Israel, Eli Magen, Eugene Merzon, Eytan Ruppin, Shlomo Vinker, Nir Giladi

## Abstract

Parkinson’s disease (PD) is characterized by progressive neurodegeneration in the autonomic and central nervous systems, manifesting with hallmark symptoms of resting tremor, bradykinesia and rigidity. The etiology of PD remains elusive, and currently available treatments do not halt disease progression.

In this study, conducted within a national health provider, we examined the impact of vaccination and medication purchase on PD occurrence and severity, using an innovative machine learning algorithm to track disease progression.

Our findings reveal a significant reduction in PD occurrence following anti-tetanus vaccination, with a time-dependent association between the elapsed time since vaccination and both the rate and progression of PD. These results are supported by evidence that antimicrobial treatments significantly alter disease severity, suggesting the actual involvement of *Clostridium Tetani* in PD pathology.

Thus, tetanus vaccination and *C. tetani* eradication could be promising strategies for preventing PD and slowing its progression, pending controlled clinical trials.

---

Parkinson’s disease (PD) is a neurodegenerative disorder affecting over six million individuals worldwide.^1^ PD is characterized by the progressive loss of neurons throughout the peripheral autonomic and central nervous systems. Current diagnosis of PD is based on the presence of resting tremor, bradykinesia, rigidity and postural response abnormalities which are tightly associated with dopaminergic loss. The etiology of PD remains poorly understood, although aging, genetic and environmental factors have been identified as risk factors. Currently, only exercise has shown an effect on disease progression and no approved treatment targets the basic mechanism of the disease or its direct cause.

Clostridia are gram-negative obligate anaerobe bacteria that are prevalent in the environment. Several members of the Clostridium species produce highly potent toxins ^2^. Notably, *C. tetani* can produce Tetanus neurotoxin, the causative agent of tetanus.^3,4^ In tetanus, the neurotoxin is internalized into signaling endosomes and transported retrogradely to the neuronal soma, interfering with the release of neurotransmitters, notably glycine and gamma-aminobutyric acid, blocking inhibitory impulses.^5^ Lacking inhibitory neurotransmission, stimulation of motor neurons increases, producing rigidity, unopposed muscle contraction and spasm.^6^

To protect from the severe risks associated with tetanus disease, the combined Tetanus and Diphtheria toxoid (TD) vaccine is routinely administered to infants, children and adolescents as part of the standard immunization programs.^7,8^ Adults usually receive anti-tetanus booster vaccination upon clinical indication, when they present in clinic with a wound susceptible to be infected with *C. tetani*, and there is no record of vaccination in the last ten years (or five years in a dirty wound at high risk for contamination). In a serologic survey performed in the United States between 1988 and 1994, fully protective levels of anti-tetanus and anti-diphtheria antibodies were detected in 91% of individuals aged 6 to 11 years but in only 47% of individuals 20 years or older ^9,10^. Interestingly, PD happens to be diagnosed at the late adult age, typically after 45, when tetanus antibody protection acquired from childhood and adolescence vaccination is no more protective. Recent studies have revealed significant differences in the gut microbiota composition between patients with Parkinson’s Disease (PD) and healthy controls ^11^. Notably, alterations were observed for Clostridia species.^12^ *C. tetani* has been isolated in the feces from adult individuals,^13^ suggesting it might be a common host of the human gut. Given its potential to cause synaptic dysfunction, clostridium neurotoxin might play a role in the neurodegenerative process responsible for PD. If a Clostridium neurotoxin contributes to the pathogenesis of PD, an inverse correlation would be expected between the administration of the tetanus toxoid vaccine and the incidence of PD.

## The study cohort

We performed a large-scale observational study in Leumit Health Services (LHS), a nationwide health organization in Israel with over 22 years of centrally maintained electronic health records (EHR). Using rigorous criteria, we selected 1446 patients who received a PD diagnosis between the ages of 45 and 75, avoiding the rare cases occurring in very young age (<45), where disease is heavily influenced by genetic or environmental factors, and older patients (>75), where non-specific motor symptoms make the clinical diagnosis of PD less reliable. We used the earliest diagnosis date or antiparkinsonian medication purchase as the index date, and selected 7230 control individuals matching in a 5:1 ratio the PD patients. Control individuals, were assigned the same index dates as their respective cases, so that the same depth of recorded EHR history was available in the two groups.

Table 1 presents the demographic and clinical comparison of the two at the index date. The age, gender and socioeconomic distribution of the two groups are very similar. In both groups 55.6% were male, with an average age of 65.7 ± 7.2. Clinical characteristics also appear to be similar, with the notable exception of smoking status, with PD patients being less likely to be smokers: Odds Ratio for current smoking (OR=0.59; P<0.001), consistent with literature.^14,15^

**Table 1.**
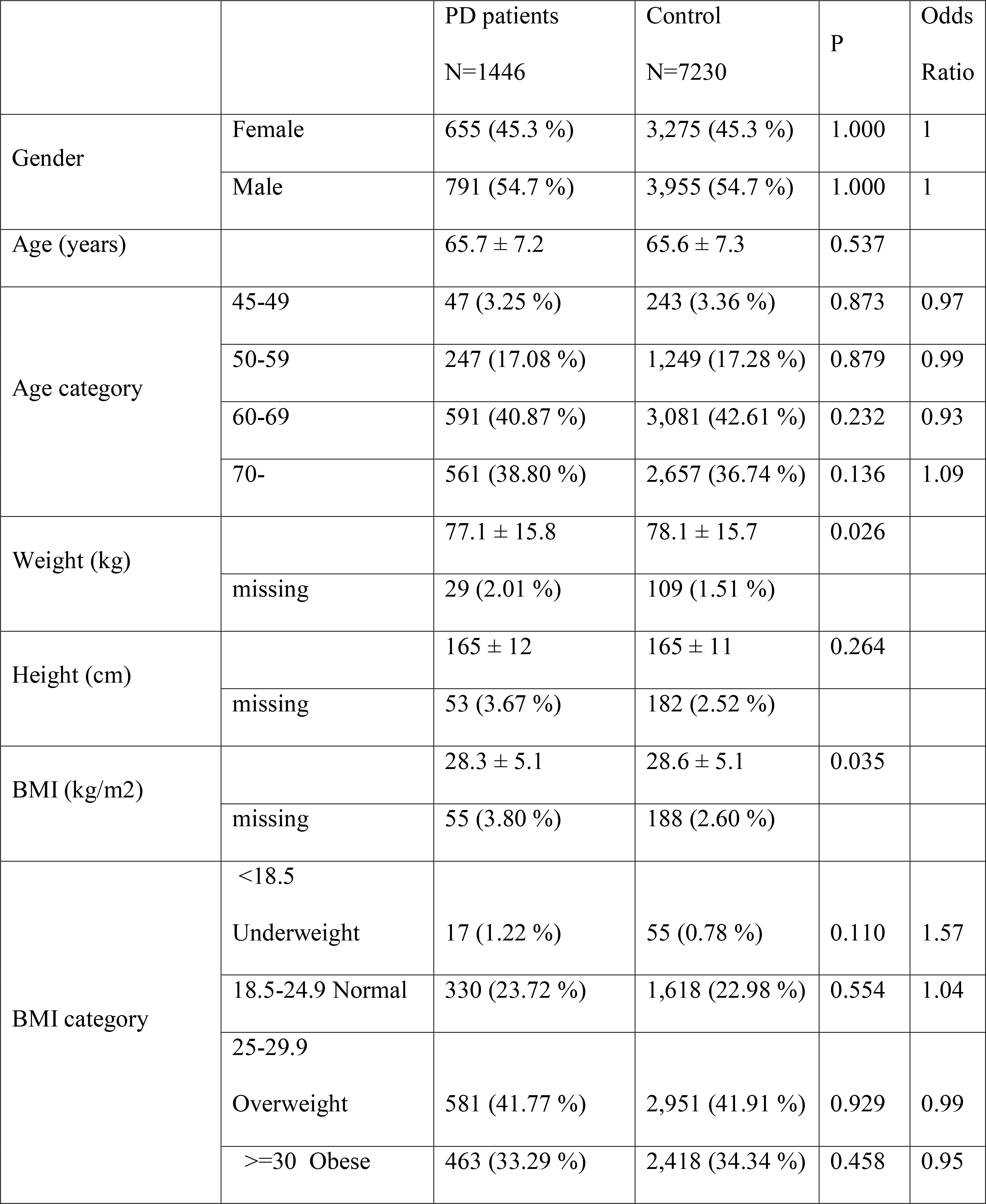
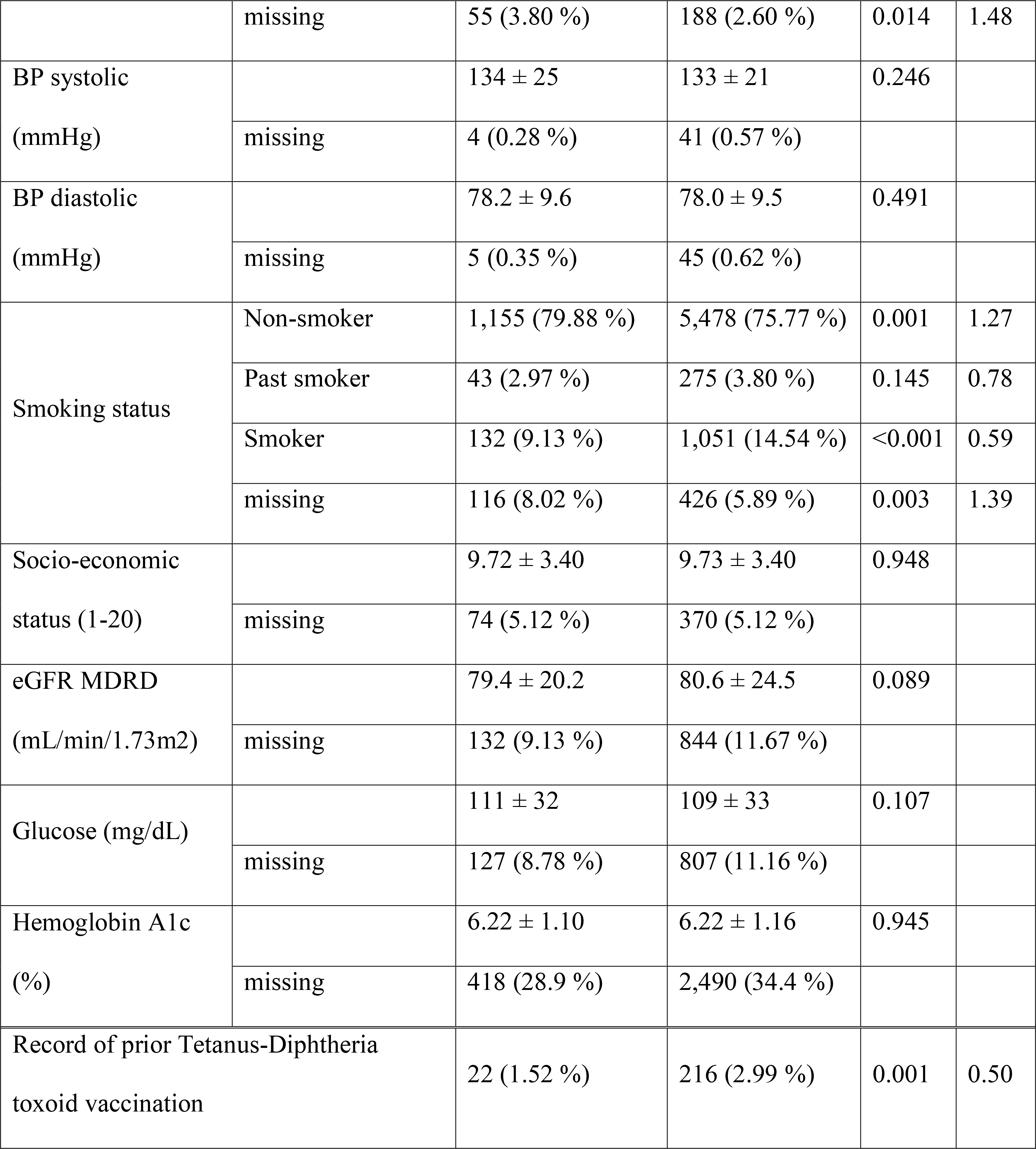
Demographic and clinical characteristics of the study cohort at index date.

## Vaccination is associated with decreased risk of PD occurrence

The last line of Table 1 displays a striking difference between the groups for TD vaccination status: only 1.64% of patients with PD had a record of vaccination before the index date, compared to 3.19% in the control group (OR=0.50, P=0.001).

A characteristic feature of a disease occurring because of waning antibody protection is a progressive increase in the disease rate with time elapsing since vaccination, as our group has shown for SARS-CoV-2.^16,17^ We therefore assessed the relationship between time elapsed since last vaccination and PD risk.

## PD risk in vaccinated patients is associated with time elapsed since vaccination

Figure 1 displays comparisons between the case and control groups according to the timing of vaccination. As the forest plot shows clearly, there is a time-dependent protection effect of the TD vaccination. The odds-ratio for PD occurring within two years after last TD vaccination is 0.00 (P=0.006), within 5 years, it is 0.17 (P=0.003). The odds increase to 0.26 between 5 to 10 years (P=0.004). Between 10- and 15-years post-vaccination, there is still a trend for protection, but the odds difference loses statistical significance. After 15 years post-vaccination the trend is for increased risk, not statistically significant.

**Figure 1.**
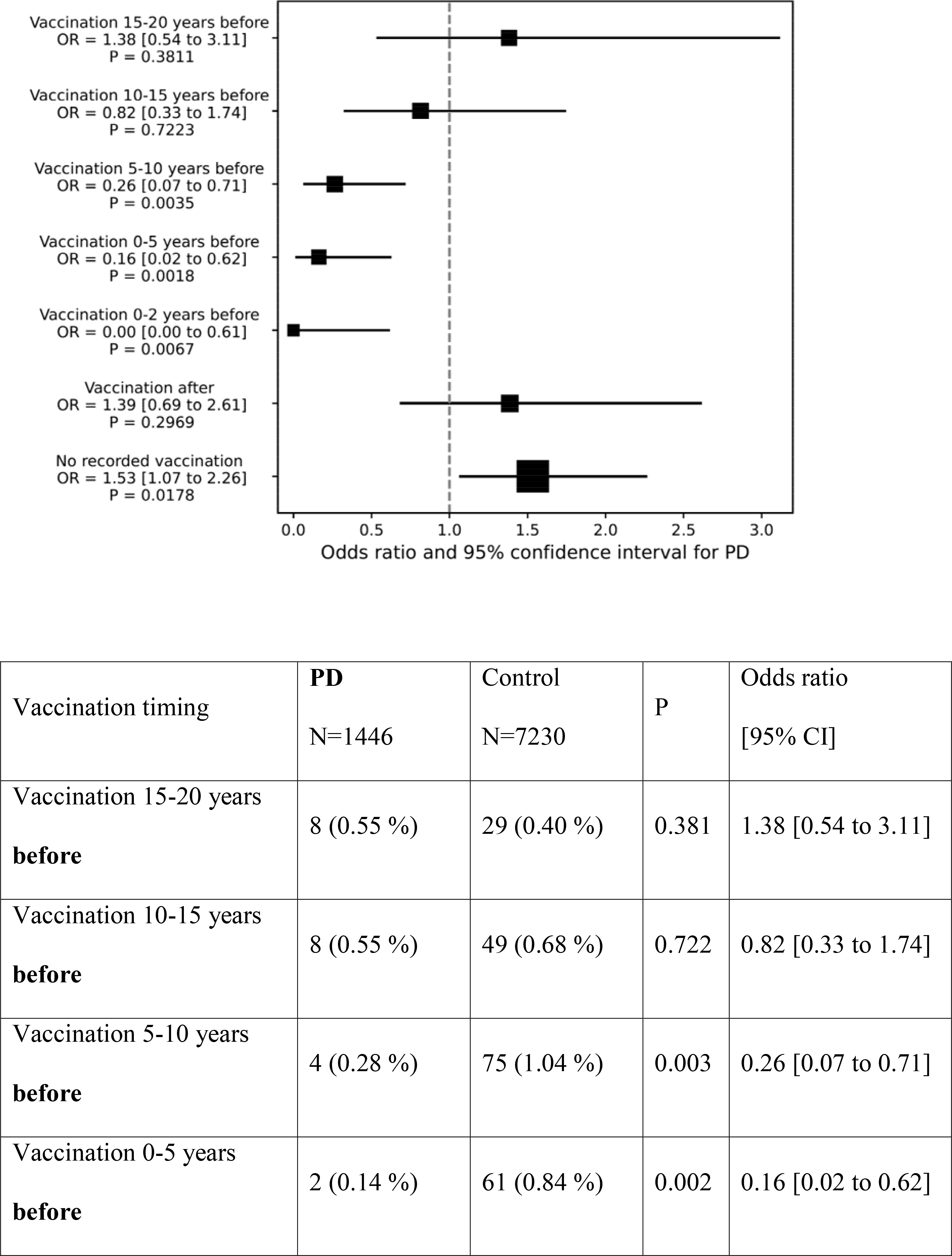
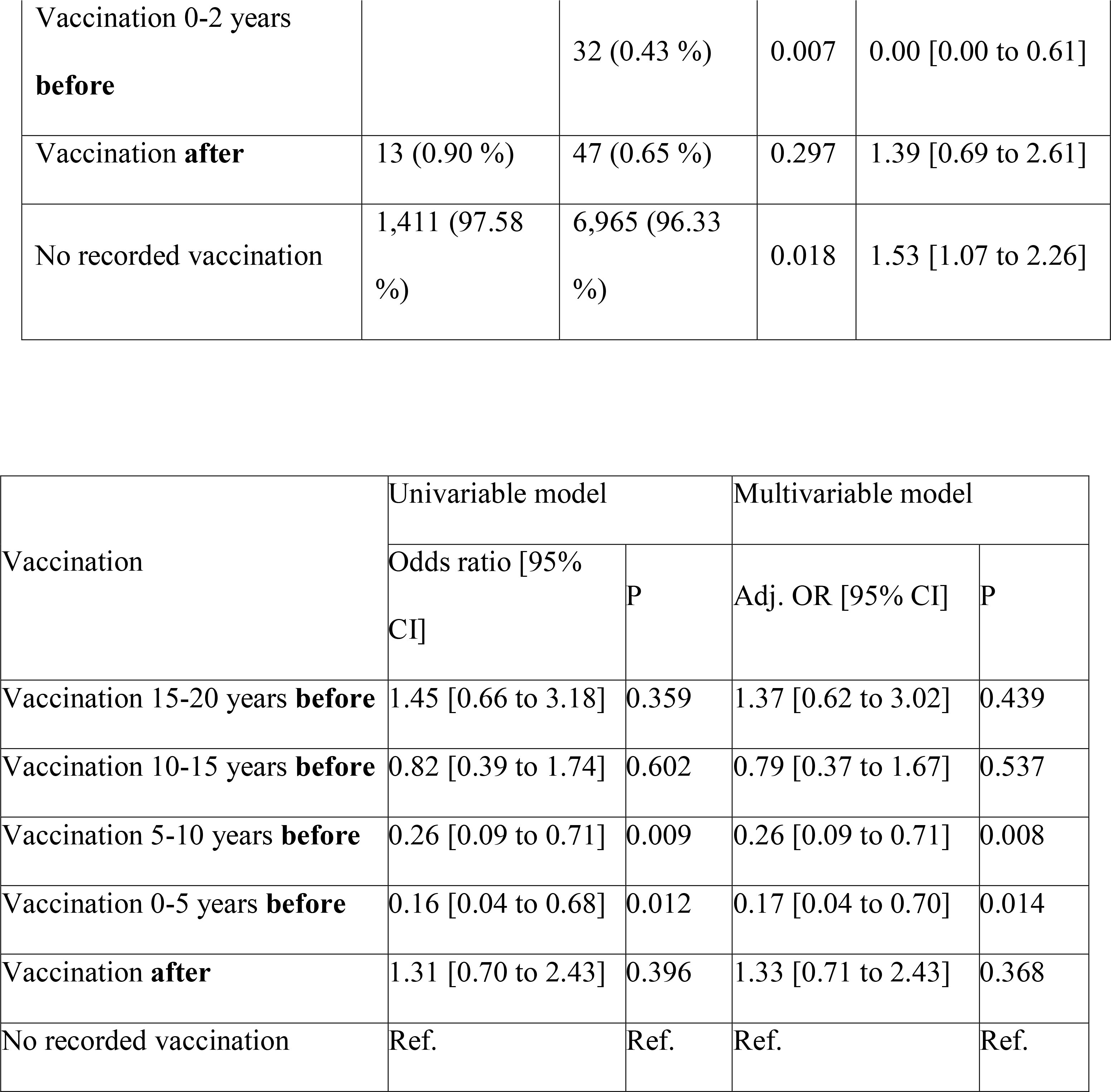
Forest plot, contingency tables, and regression models of Tetanus Diphteria (TD) vaccinations according to its timing with regards to the index date

If not getting a TD vaccination was to be a confounder of some early symptoms of PD (e.g. people with early signs of PD, before diagnosis, may decrease their activity and hence reduce opportunities to get wounded, and subsequently get vaccination), then one would expect that this pattern would continue and even amplify after the index date. But this does not occur: instead, we observe an opposite trend, of increased odds for PD among those who got a vaccine after the index date (OR=1.33, P=0.39). This reversing time relationship strongly indicates that lack of TD vaccination is unlikely to be a confounder for early PD.

Having shown that PD occurrence is highly associated with time elapsed since last TD vaccination, we proceeded to verify, in the few patients who were either diagnosed with PD after vaccination, or were vaccinated once PD was diagnosed, whether disease course was affected by vaccination.

## Vaccinated PD patients have slower rates of disease progression

In order to follow the disease course, we developed a method to accurately assess disease severity during the follow-up period. Parkinson’s disease is characterized by progressive course of motor and functional deterioration as shown by Hoehn & Yahr’s clinical staging of the laterality and axial symptoms^18^ and by the Unified Parkinson’s Disease Rating Scale (UPDRS).^19^ Symptoms are typically treated by antiparkinsonian medications that are increased over time as the disease progresses in dosage and potency. Having records of medications purchased by each patient, we could use the annual medication consumption to train a machine learning model, that would assess disease severity for each patient and year of disease. For this purpose, we trained a gradient boosting model by cross-validation folds, utilizing yearly medication consumption per catalog entry as training variables, together with the gender of the patient (since disease course is affected by gender), to estimate time elapsed since disease onset, set as the target variable. The output of the model is a PD severity score expressed in a scale analogous to years of disease (e.g. a patient with a severity score of 6 in a given year has purchased medications that are typically used by a PD patient in the 6th year of the disease). Using this model, we computed severity scores for 8,793 PD patient years of disease, of which 201 were from patients with prior vaccination record.

Figure 2A displays a kernel density plot of these scores vs. the actual disease duration, along with a linear regression line. The calculated severity scores correlate nicely with the actual year of the disease. The Pearson correlation coefficient, is 0.443, with a P-value under 10^-200^. Of note, a quadratic regression model (a polynomial model of degree 2) was slightly more informative, exhibiting a lower Akaike Information Criterion (AIC) and a higher R^2^ compared to the linear model (see Fig. 2B).

**Figure 2.**
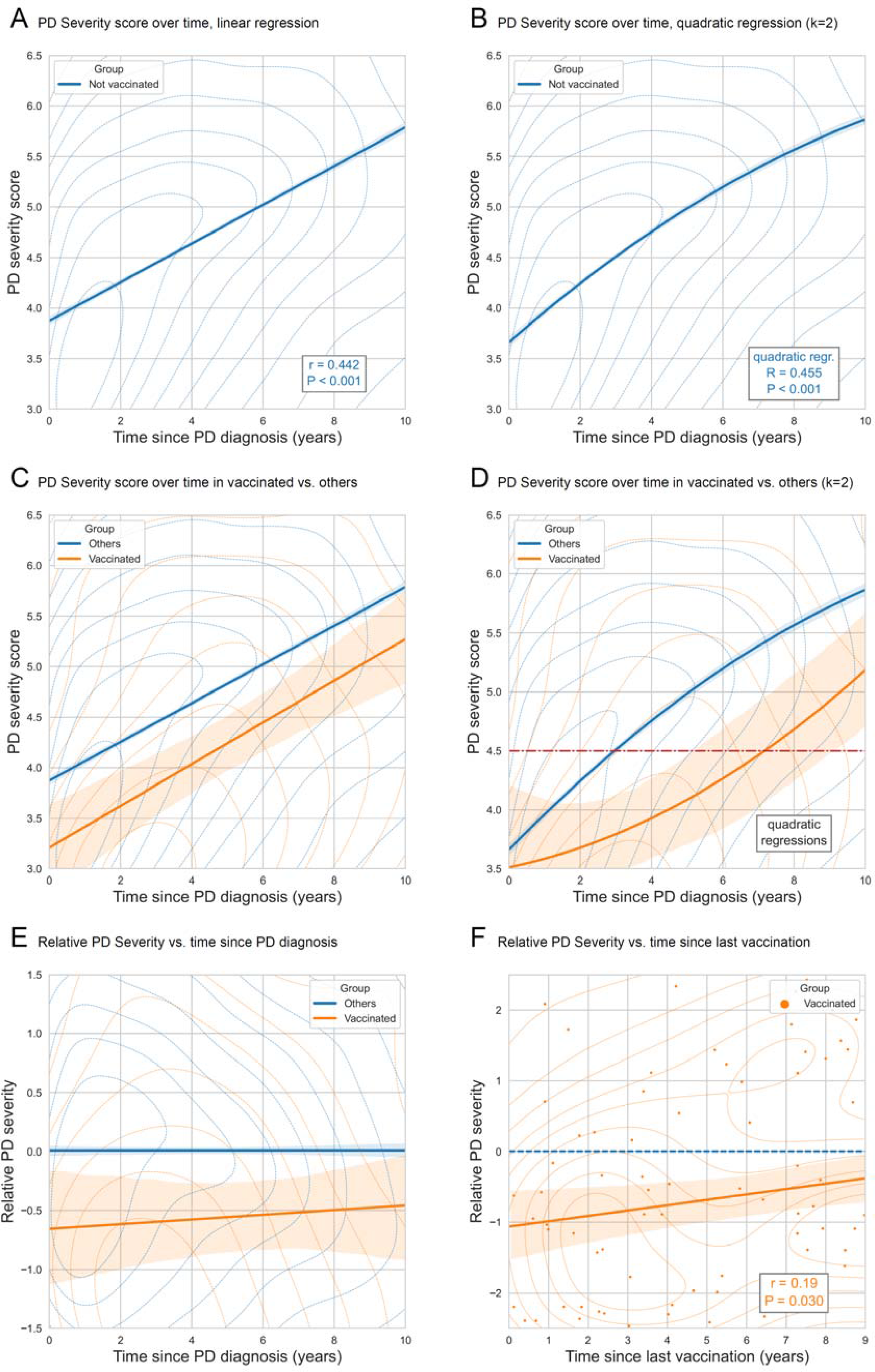
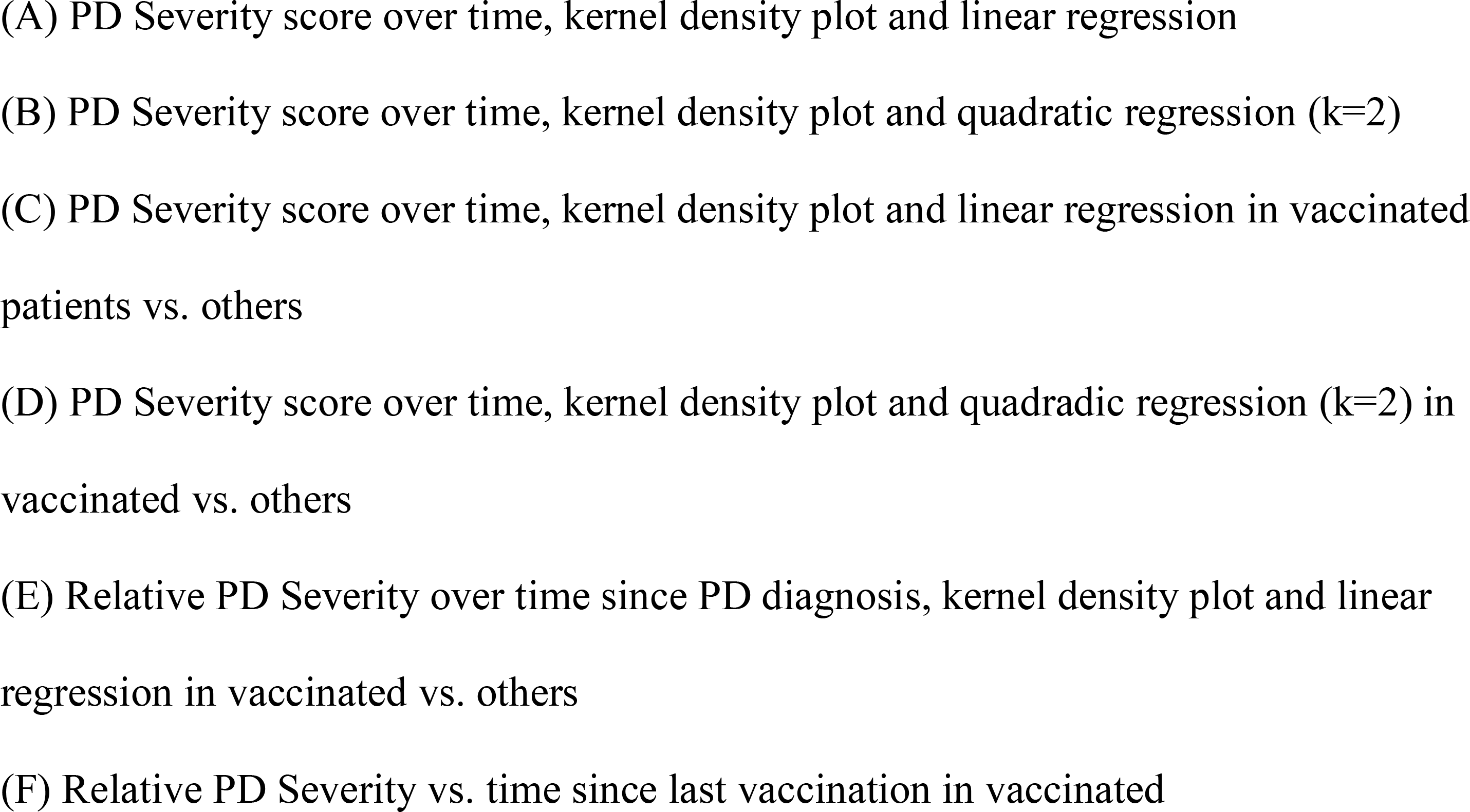
Severity kernel density plots and regression analyses

Figure 2C displays the severity scores over time, according to vaccination status. Disease severity in vaccinated patients (orange curve) was significantly lower than in non-vaccinated patients. Linear regression models show vaccination status is significantly associated with reduced severity, both in a univariable model (P<0.001, Table S1), and in a multivariable model adjusted for age, gender and smoking status (P<0.001, Table S2). Here again the quadratic regression is more informative (Table S3), so we use a quadratic model to plot disease severity according to time elapsed since PD diagnosis: Figure 2D shows that, even though disease severity was roughly similar at disease onset, disease progression was significantly slower in the vaccinated group. Their regression curve shows that the average severity in vaccinated patients at the seventh year of the disease (red line) was similar to the average severity score of unvaccinated patients who were at the third year of the disease.

We see here, that even among PD patients, disease course was less severe post-vaccination than in PD patients with no prior vaccination record.

## Disease progression is associated with time elapsed since vaccination

We have shown that PD progresses more slowly in patients who have been vaccinated than in those who did not. A further indication that TD vaccine affects PD progression would be to find, focusing on the vaccinated patients, an effect that is more pronounced when closer to the date of vaccination, even after correcting for time since diagnosis. For this analysis, which requires comparison of patients who have experienced varying durations of the disease, we calculated “relative PD severity scores”, which are obtained by subtracting the mean severity score of patients at the same disease year from the severity scores.

Figure 2E displays these relative PD severity scores according to time since diagnosis, divided by vaccination status, showing once again that vaccinated individuals have lower disease severity (P<0.001, Table S4). Figure 2F follows the relative disease severity of vaccinated patients according to time elapsed since last vaccination. The relative disease severity decrease is correlated with time elapsed since vaccination (r=0.19, P=0.03, Tables S5 and S6 for crude and adjusted regression analyses).

## Disease severity is affected by antimicrobial treatments

If *C. tetani* present in the patients’ microbiome are actually involved in PD pathology, then PD disease severity should be affected by antimicrobial treatments that kill these bacteria. Having calculated PD disease severity scores, we can use them to assess whether antimicrobial courses are actually associated with changes in disease severity. For this analysis, we identified forty classes of medications for which we observed varying consumption rates between PD patients and control patients in the years prior to the index date, which is a potential signal for an effect on disease occurrence.^20^ For each of these classes, and for each year of disease in a PD patient, we calculated a variable that reflects whether medications of the class were purchased by the patient over the three preceding years. Starting with a multivariable regression model that includes age, gender, and smoking status, we employed a stepwise approach to incrementally select the ten medication classes most strongly associated with the relative PD severity scores. Table 2 presents the results of this regression. Not surprisingly, the tetanus toxoid vaccine had the strongest effect on disease severity, purchase of the vaccine in the preceding three years significantly reduced the relative disease severity (-1.11, P=0.0003).

**Table 2.** Multivariable regression model examining the impact of medication purchased in the three preceding years on relative PD disease severity.

| ATC code |  | Estimate | 95% Confidence Interval | P-value |
| --- | --- | --- | --- | --- |
|  | Age | -0.00 | [-0.01 to 0.00] | 0.157 |
|  | Smoker | 0.18 | [0.05 to 0.32] | 0.006 |
| J01MA01 | Ofloxacin | -0.45 | [-0.55 to -0.34] | $7.08 \cdot 10^{-17}$ |
| A06AD15 | Macrogol | 0.34 | [0.23 to 0.44] | $3.09 \cdot 10^{-10}$ |
| J01FA01 | Erythromycin | -0.74 | [-1.12 to -0.35] | 0.0002 |
| A06AC01 | Ispaghula (Psyllium) | -0.42 | [-0.65 to -0.19] | 0.0004 |
| J07AM51 | Tetanus-Toxoid vaccine | -1.11 | [-1.71 to -0.51] | 0.0003 |
| J01CE08 | Benzathine<br>Benzylpenicillin | -0.69 | [-1.15 to -0.24] | 0.0028 |
| J01DB05 | Cefadroxil | -1.02 | [-1.70 to -0.35] | 0.0029 |
| J01FF01 | Clindamycin | 0.40 | [0.15 to 0.64] | 0.0017 |
| J01FA06 | Roxithromycin | -0.13 | [-0.23 to -0.04] | 0.0076 |
| J01MA12 | Levofloxacin | -0.22 | [-0.41 to -0.03] | 0.0263 |

Several antibiotic agents displayed a substantial and significant effect in reducing disease severity: benzathine penicillin, which is an intramuscular formulation of penicillin, displayed a significant reducing effect on disease severity (-0.69, P=0.003). Benzathine penicillin is the classical treatment for tetanus,^21^ its sustained release formulation is likely to deplete the clostridial population producing the toxin, explaining its beneficial effect on disease severity. Likewise, Cefadroxil, a first generation cephalosporine structurally close to penicillin displayed a significant decreasing effect on disease severity (-1.02, P=0.003). Two macrolide antibiotics also displayed a beneficial effect on disease severity: erythromycin (-0.74, P=0.0002) and roxithromycin (-0.13, P=0.008), as well as two compounds of the fluoroquinolone family: ofloxacin (-0.45, P<0.001), and levofloxacin (-0.22, P=0.026).

On the other hand, Clindamycin was associated with a significant and substantial increase in disease severity (+0.40, P=0.002). Clindamycin is primarily used to treat infections caused by susceptible anaerobic bacteria, but it is notorious for causing *Clostridium difficile* colitis, a dangerous condition in which Clostridium bacteria, inherently resistant to clindamycin, colonize the human colon.^22^ The observed effect of increasing severity of PD is consistent with a competitive advantage granted by this antibiotic to clostridia, over susceptible anaerobes present in the microbiome, enabling proliferation of *C. tetani* in a manner similar to *C. difficile* proliferation following Clindamycin treatment.

Interestingly, we also observed strong apposing effects for two laxative agents commonly used to treat constipation in PD patients. Macrogol (polyethylene glycol), was significantly associated with increased disease severity (+0.34, P<0.001), while ispaghula (Psyllium), a dietary fiber, was associated with decreased disease severity (-0.42, P=0.0004). Since these two compounds primary affect the gut environment, these opposite effects are strongly suggestive of a pivotal role played by an organism which resides in the intestinal microbiome.

We tested whether a dose-response relationship could be detected, by looking for a correlation between the number of purchases by each patient during the three preceding years and the relative PD severity score. Figure 3 display scatter plots and correlation analyses for the identified compounds. For most compounds we observe a significant dose-response relationship strengthening the direction of the association detected by the regression. Since the regression was based only on a binary variable for each compound, and did not account for the number of purchases, a significant dose-response relationship provides an independent corroboration of the effect of these medications.

**Fig. 3.**
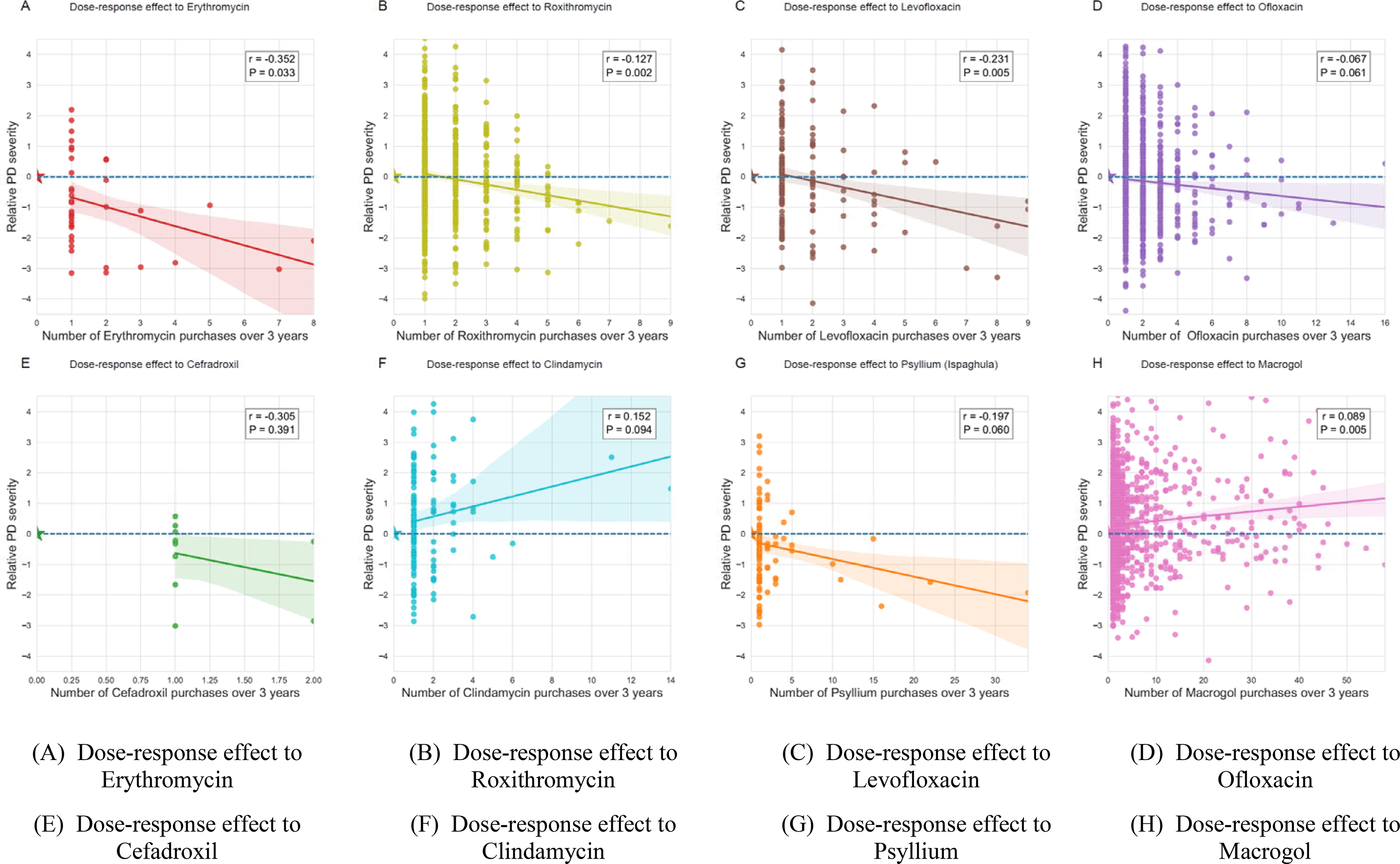
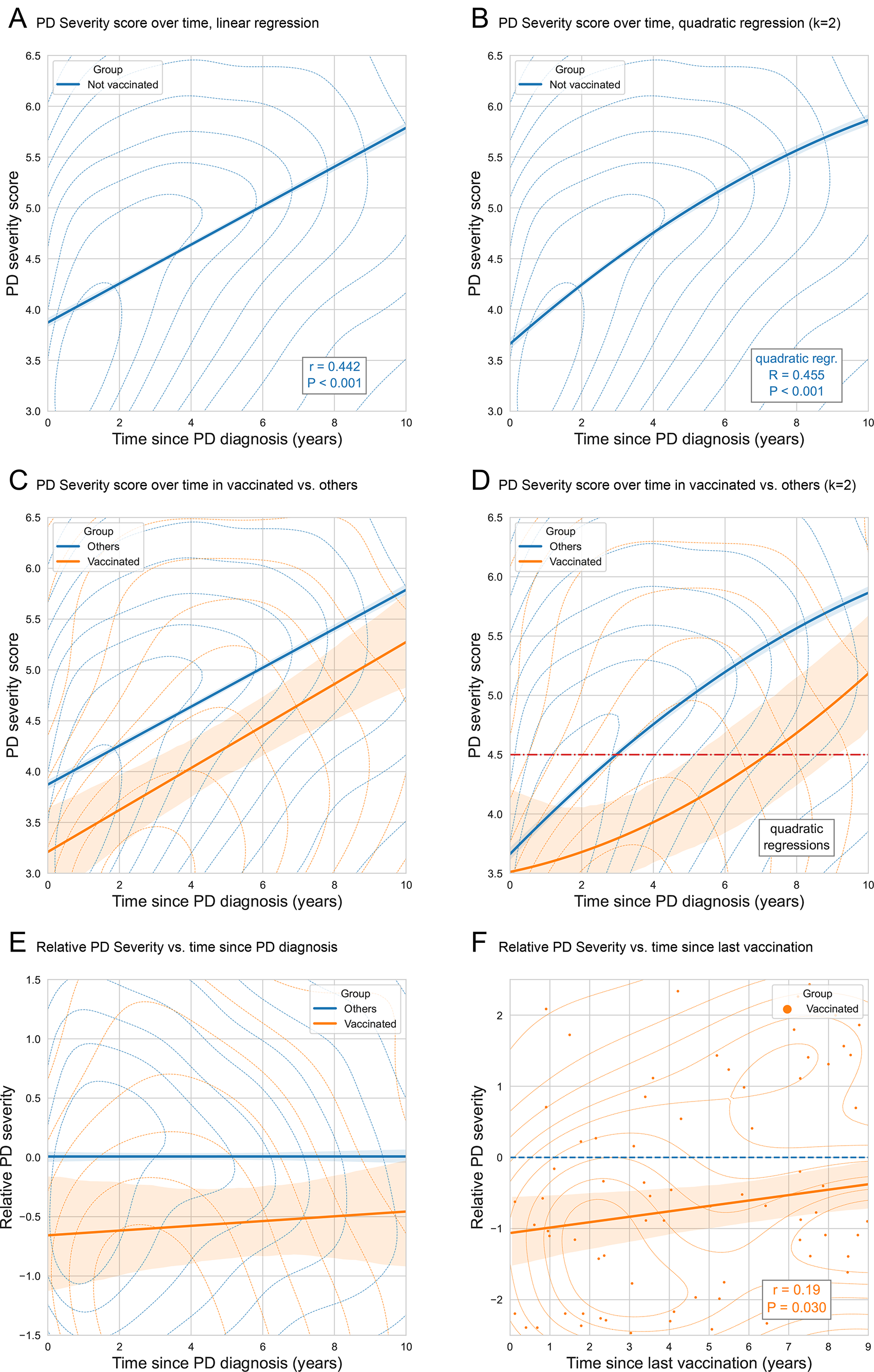
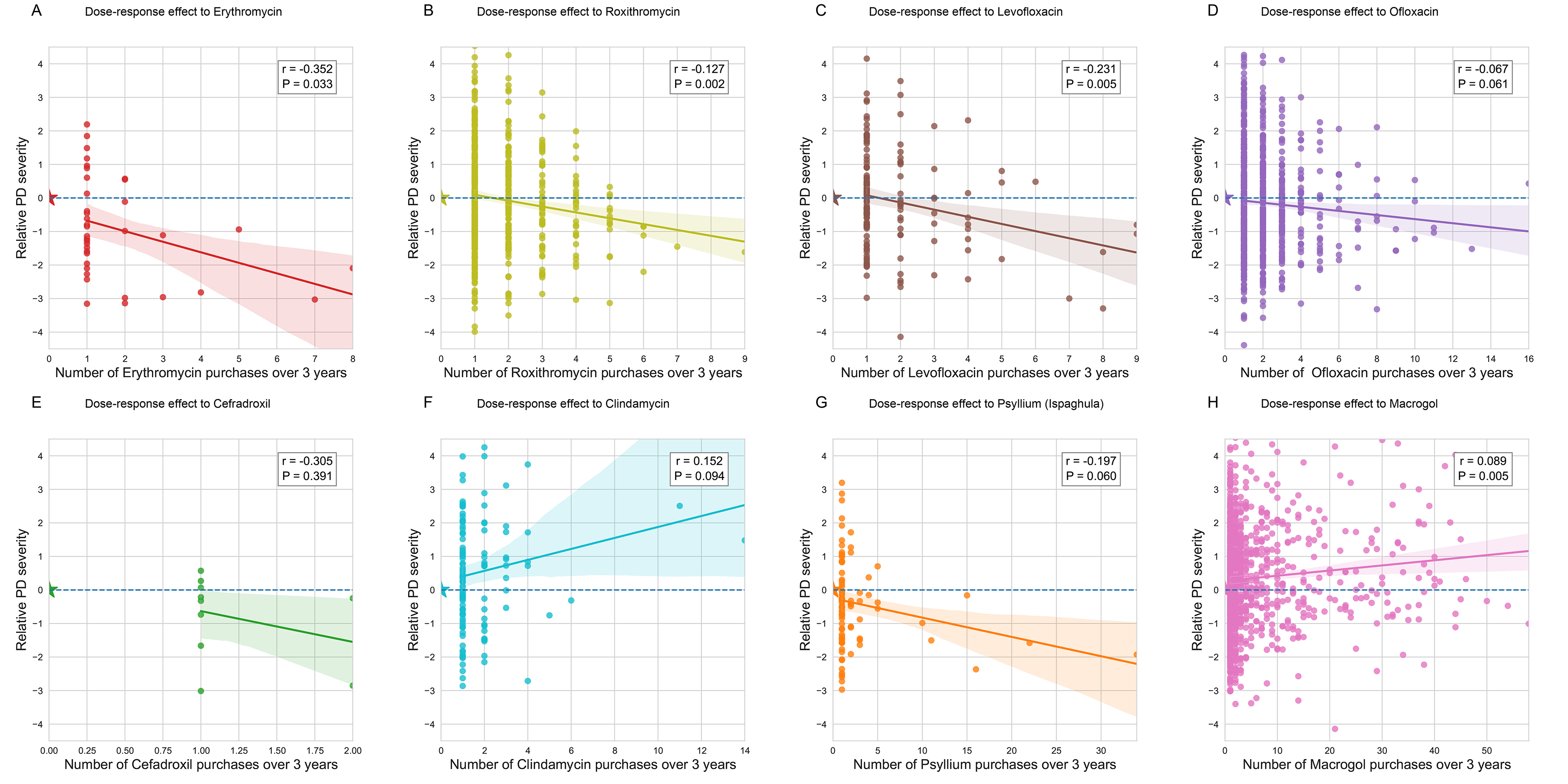
Dose-response effect associated with purchase of selected antimicrobial agents and food supplements

## Discussion

This large-scale, real-life population study provides compelling evidence for the protective effect of diphtheria-tetanus (DT) toxoid vaccination on PD risk and its rate of progression. The significant associations observed between time elapsed since vaccination and both PD occurrence and rate of disease progression are all supporting the interpretation that the anti-tetanus vaccine provides protection against PD, and that this protection wanes over time. In addition, disease progression is significantly associated, in a dose-dependent manner, to antibiotics that affect clostridium growth negatively (beta-lactam, macrolides, fluroquinolones), or positively (clindamycin), strongly supporting the involvement of actual clostridium bacteria as the causative agent of disease.

Clostridia are known to reside in the microbiome, and several studies have reported differential prevalence of clostridial species in the gut microbiome of PD patients.^11,12^ There is strong indication that *C. tetani* is a common host of the human microbiome: a study performed in Vietnam incidentally reported *C. tetani* presence in the feces of 42% of control subjects.^13^ *C. tetani* are also known to be present in the saliva, and contamination of a wound with saliva in an unvaccinated individual is indication for treatment with tetanus immunoglobulin.^23^ *C. Tetani* spores are prevalent in the human environment, extremely resistant: properties that should allow this organism to progressively colonize the human flora. *C. tetani* toxin secreted in the gut would primarily affect the mesenteric neurons that regulate peristalsis. Remarkably, impairment of the autonomic nervous system of the gut, manifested as constipation and bloating, happens to be a very common prodromal non-motor symptom of PD.^24^

The subsequent spreading of *C. tetani* to other microbiome niches would explain how the pathology of PD is further observed in multiple foci. *C. tetani*, present in the nasopharynx would damage by its neurotoxin the olfactory neurons, affecting smell.^25^ Once significant damage is incurred to dopamine secreting neurons within the basal ganglia via retrograde axonal transport,^26^ the disease manifests by the characteristic movement disturbances of PD,^27^ while nonspecific neurodegeneration presents as depression and dementia.^28^ In the mouth, *C. tetani* would affect the innervation of the salivary glands (causing drooling); invasion of the sebaceous glands would dysregulate sweating, a common PD symptom.

The oropharynx and the paranasal sinuses, which lie in close proximity to the CNS, are the primary sites we suspect to harbor *C. tetani* in PD. In these sensitive sites, even trace amounts of the extremely potent tetanus neurotoxin would be expected to cause over time synaptic dysfunction and subsequent damage to neighboring neuronal tissue. The α- synuclein aggregates that are the hallmark of PD could hence be a marker of synaptic dysfunction triggered by the toxin rather than the original cause of neurodegeneration.

Given its prevalence in the environment, presence of *C. tetani* in the microbiome, even after loss of the humoral response that could neutralize its toxin, is certainly not the sole factor of PD occurrence. Genetic and environmental factors, as well as lifestyle and comorbidities, likely influence the ability of *C. tetani* to colonize susceptible niches and harm adjacent neuronal tissue. Interestingly, cigarette smoking has been shown to inhibit Clostridium growth,^29^ hence, mere inhibition of C*. tetani* proliferation can explain the decreased risk observed for PD in smokers.^14,15^

In conclusion, the findings presented in this study strongly support a causative role played by *C. tetani* in the pathological process leading to diffuse and progressive neurodegeneration observed in PD. Based on solid epidemiological data, we propose here a new paradigm that offers a mechanistic explanation of PD pathogenesis and its wide diversity of symptoms progression, in the form of neurotoxin mediated damage caused by

### *C. tetani* colonizing the microbiome at sensitive sites

This study suggests that vaccination against tetanus neurotoxin (active or passive), possibly combined with treatments aimed at eradicating *C. tetani* from body reservoirs, could offer promising avenues to prevent Parkinson’s Disease (PD) occurrence and slow disease progression. Our findings make a strong case for a large-scale prospective randomized controlled trial to test and validate the protective effect of anti-tetanus vaccination on PD progression in early and advanced stages of PD, as well as in subjects at risk. Additionally, further research should explore factors affecting *C. tetani* proliferation and colonization potential, particularly in relation to medication, diet, food supplements, hygiene products, and lifestyle habits.

## Methods

### Ethics Statement

The Institutional Review Board (IRB) of Leumit Health Services gave ethical approval for this work (LEU-0001-24), with a waiver of informed consent, since data were analyzed retrospectively and anonymously.

### Study design

This research was conducted as an observational cohort study in Leumit Health Services (LHS), one of the four national health providers in Israel, providing comprehensive healthcare services to approximately 720,000 members. All Israeli citizens are entitled for comprehensive health insurance and receive a standardized package of health services and medications, as defined by the national “Health Basket” committee. LHS operates a centralized electronic health records (EHR) system, with over two decades of meticulously maintained information on patient demographics, medical diagnoses, healthcare encounters, laboratory test results, and records of prescribed and purchased medications. Diagnoses are documented during medical encounters by the treating physicians using the International Classification of Diseases, Ninth Revision (ICD-9).

Diagnoses can be marked as chronic when they pertain to a chronic condition, and these can be updated or closed by the treating physicians during subsequent patient encounters. The reliability of these chronic diagnosis records in our registry has been previously validated, demonstrating high accuracy.^16^

Eligibility for inclusion in the study was defined as any past or current LHS member with at least five years of LHS membership between years 2003 and 2023. Data extraction was carried out from the LHS central data warehouse in February 2024, encompassing diagnoses, results of laboratory tests, and medication purchases recorded up to December 31, 2023.

### Cohort definition

The study cohort comprises eligible PD patients diagnosed for the first time between ages 45 and 75, alongside a control group matched at a 5:1 ratio, of individuals with no documented PD. PD patients were identified by the presence of ICD-9 coded 332 diagnosis “Parkinson’s Disease” in the EHR, if recorded by a neurologist or a movement disorder specialist, or recorded by any physician, if accompanied with purchase of Antiparkinsonian medications (listed in Table S7) over a period of more than six months. In order to avoid confusion with overlapping conditions, individuals with a diagnosis indicative of secondary parkinsonism, schizophrenia, pituitary adenoma, restless legs syndrome, or cerebrovascular accident were excluded from the study (diagnoses listed in Table S8), as well of individuals with prior purchase of an antipsychotic medication susceptible to induce parkinsonism (medications listed in Table S9).

Controls were precisely matched to the PD patients based on gender, socio-economic status category, and the year of initial enrollment in Leumit Health Services (LHS). For each PD patient, five control individuals were chosen who met these matching criteria and whose birth dates were closest to that of the PD patient, ensuring no individual was duplicated within the cohort.

### Diphtheria-Tetanus toxoid vaccinations

We looked for anti-tetanus vaccinations in the medical history of patients from the study cohort, using both vaccine purchase recorded by LHS pharmacies, and vaccine administration records documented in the EHRs. Most anti-tetanus vaccinations performed in LHS for patients in the cohort were with DT IMOVAX from Sanofi-Pasteur, adult dose, 0.5ml.

### Data preparation

Data were extracted from electronic health records and prepared for analyses using scripts developed by Leumit Research Institute in Python 3.11 with Pandas library and T-SQL queries. Prior to analysis, patients’ data were deidentified and the patient ID number replaced with an identifier internal to the study.

### Statistical analyses

Statistical analyses were performed in R version 4.3. Unless specified otherwise, Fisher’s exact test was used to compare categorical variables and the two-tailed two samples t-test to compare continuous variables across groups. Conditional logistic models were fit to assess the association of vaccination timing and covariates across matched groups. Linear regression models were fit to assess the relationship between linear variables such as disease severity and relative disease severity and explanatory variables. Pearson correlation analyses were performed to assess dose-response relationship. Graphs of this study were produced in Python with *seaborn* and *matplotlib* libraries.

A.I. and E.Me had access to all the original data and serve as guarantor of the study integrity.

### Machine Learning Model

Gradient boosting model for predicting disease activity score was built with *lightgbm* python library following exploration of model parameters space performed with *FLAML/AutoML* with 5-fold cross-validation. The predicting features of the model are provided as Supplementary Table S10.

## Data Availability

Access to patients data is limited to researchers approved by the Institutional Review Board.

## End Notes

### Acknowledgements

We thank Alejandro A. Schäffer for his assistance in preparation of this manuscript. The content of this publication does not necessarily reflect the views or policies of the Department of Health and Human Services, nor does mention of trade names, commercial products, or organizations imply endorsement by the US Government.

### Author contributions

Conceptualization: AI, EMa Methodology: AI, EMe Investigation: AI, NG

Writing – original draft: AI, NG

Writing – review & editing: AI, EMa, EMe, ER, SV, NG

### Supplementary Materials

Supplementary Information is available for this paper.

### Materials & Correspondence

Correspondence and requests for materials should be addressed to AI.

