## Supplementary Material for "Anti-tetanus vaccination is associated with reduced occurrence and slower progression of Parkinson’s disease"

**The PDF file includes:**

Tables S1 to S10

Table S1.

Linear regression of severity score vs. time since PD diagnosis

|  | Coef | 95% Confidence Interval | P. Value |
| --- | --- | --- | --- |
| (Intercept) | 3.87 | [3.82 to 3.92] | 0.00000 |
| vaccinated | -0.59 | [-0.87 to -0.31] | 0.00005 |
| Year since diagnosis | 0.19 | [0.18 to 0.20] | 0.00000 |

Table S2.

Linear regression of severity score vs. time since PD diagnosis adjusted for age, gender, and smoking status

|  | Coef | 95% Confidence Interval | P. Value |
| --- | --- | --- | --- |
| (Intercept) | 3.940 | [3.63 to 4.25] | 0.00000 |
| vaccinated | -0.530 | [-0.81 to -0.24] | 0.00029 |
| Year since diagnosis | 0.190 | [0.18 to 0.20] | 0.00000 |
| age | 0.000 | [-0.01 to 0.00] | 0.19372 |
| Gender Female | 0.270 | [0.20 to 0.34] | 0.00000 |
| smoker | 0.180 | [0.05 to 0.32] | 0.00878 |

**AIC 8249.830**

Table S3.

Linear regression of severity score vs. time since PD diagnosis (quadratic) adjusted for age, gender, and smoking status

|  | Coef. | [CI] | Pr(>\|t\|) |
| --- | --- | --- | --- |
| (Intercept) | 3.760 | [3.46 to 4.07] | 0.00000 |
| vaccinated | -0.540 | [-0.82 to -0.26] | 0.00016 |
| Year since diagnosis | 0.310 | [0.29 to 0.33] | 0.00000 |
| I(Year since diagnosis^2) | -0.010 | [-0.01 to -0.01] | 0.00000 |
| age | 0.000 | [-0.01 to 0.00] | 0.12073 |
| Gender Female | 0.280 | [0.21 to 0.34] | 0.00000 |
| smoker | 0.200 | [0.07 to 0.34] | 0.00290 |

**AIC 8132.973**

Table S4.

### Linear regression of relative PD severity vs. vaccination status and time elapsed since PD diagnosis

|  | Coef | [CI] | Pr(>\|t\|) |
| --- | --- | --- | --- |
| (Intercept) | 0.010 | [-0.04 to 0.06] | 0.80900 |
| vaccinated | -0.560 | [-0.84 to -0.28] | 0.00009 |
| Year since vaccination | 0.000 | [-0.01 to 0.01] | 0.93520 |

**AIC 8057.771**

Table S5.

### Linear regression of relative PD severity in vaccinated according to time elapsed since vaccination

|  | Coef | [CI] | Pr(>\|t\|) |
| --- | --- | --- | --- |
| (Intercept) | -1.060 | [-1.61 to -0.51] | 0.00024 |
| Year since vaccination | 0.070 | [0.01 to 0.14] | 0.03447 |

**AIC 129.9**

Table S6.

### Linear regression of relative PD severity in vaccinated according to time elapsed since vaccination adjusted for covariates

|  | Coef | [CI] | Pr(>\|t\|) |
| --- | --- | --- | --- |
| (Intercept) | 9.060 | [5.33 to 12.80] | 0.00001 |
| Year since vaccination | 0.070 | [0.00 to 0.13] | 0.05090 |
| Year since diagnosis | 0.130 | [0.05 to 0.21] | 0.00147 |
| age | -0.150 | [-0.20 to -0.09] | 0.00000 |
| Gender Female | -0.850 | [-1.60 to -0.10] | 0.02805 |
| smoker | -2.650 | [-4.17 to -1.12] | 0.00091 |

**AIC 110.15**

Table S7.

List of medications used to determine Parkinson’s Disease

| N04AA04 | Procyclidine |
| --- | --- |
| N04BA02 | Levodopa and Decarbo |
| N04BA03 | Levodopa, Decarboxyl |
| N04BA07 | foslevodopa and deca |
| N04BB01 | Amantadine |
| N04BC02 | Pergolide |
| N04BC04 | Ropinirole |
| N04BC05 | Pramipexole |
| N04BC07 | Apomorphine |
| N04BD01 | Selegiline |
| N04BD02 | Rasagiline |
| N04BD03 | Safinamide |
| N04BX02 | Entacapone |

Table S8.

List of diagnoses excluding eligibility in the cohort

| **ICD9 code** | **Diagnosis description** |
| --- | --- |
| 225.11 | ADENOMA OF HYPOPHYSE |
| 227.3 | ADENOMA OF HYPOPHYSE-BENIGN NEOPLASM OF PITUITARY GLAND AND CRANIOPHARYNGEAL DUCT |
| 253.12 | PROLACTINOMA |
| 253.120 | MICROADENOMA OF HYPOPHYSE |
| 253.121 | MACROADENOMA OF HYPOPHYSE |
| 295 | SCHIZOPHRENIC DISORDERS |
| 295.0 | SIMPLE TYPE SCHIZOPHRENIA |
| 295.00 | SIMPLE TYPE SCHIZOPHRENIA, UNSPECIFIED STATE |
| 295.01 | SIMPLE TYPE SCHIZOPHRENIA, SUBCHRONIC STATE |
| 295.02 | SIMPLE TYPE SCHIZOPHRENIA, CHRONIC STATE |
| 295.03 | SIMPLE TYPE SCHIZOPHRENIA, SUBCHRONIC STATE WITH ACUTE EXACERBATION |
| 295.04 | SIMPLE TYPE SCHIZOPHRENIA, CHRONIC STATE WITH ACUTE EXACERBATION |
| 295.05 | SIMPLE TYPE SCHIZOPHRENIA, IN REMISSION |
| 295.1 | DISORGANIZED TYPE SCHIZOPHRENIA |
| 295.10 | DISORGANIZED TYPE SCHIZOPHRENIA, UNSPECIFIED STATE |
| 295.11 | DISORGANIZED TYPE SCHIZOPHRENIA, SUBCHRONIC STATE |
| 295.12 | DISORGANIZED TYPE SCHIZOPHRENIA, CHRONIC STATE |
| 295.13 | DISORGANIZED TYPE SCHIZOPHRENIA, SUBCHRONIC STATE WITH ACUTE EXACERBATION |
| 295.14 | DISORGANIZED TYPE SCHIZOPHRENIA, CHRONIC STATE WITH ACUTE EXACERBATION |
| 295.15 | DISORGANIZED TYPE SCHIZOPHRENIA, IN REMISSION |
| 295.2 | CATATONIC TYPE SCHIZOPHRENIA |
| 295.20 | CATATONIC TYPE SCHIZOPHRENIA, UNSPECIFIED STATE |
| 295.21 | CATATONIC TYPE SCHIZOPHRENIA, SUBCHRONIC STATE |
| 295.22 | CATATONIC TYPE SCHIZOPHRENIA, CHRONIC STATE |
| 295.24 | CATATONIC TYPE SCHIZOPHRENIA, CHRONIC STATE WITH ACUTE EXACERBATION |
| 295.25 | CATATONIC TYPE SCHIZOPHRENIA, IN REMISSION |
| 295.3 | PARANOID TYPE SCHIZOPHRENIA |
| 295.30 | PARANOID TYPE SCHIZOPHRENIA, UNSPECIFIED STATE |
| 295.31 | PARANOID TYPE SCHIZOPHRENIA, SUBCHRONIC STATE |
| 295.32 | PARANOID TYPE SCHIZOPHRENIA, CHRONIC STATE |
| 295.33 | PARANOID TYPE SCHIZOPHRENIA, SUBCHRONIC STATE WITH ACUTE EXACERBATION |
| 295.34 | PARANOID TYPE SCHIZOPHRENIA, CHRONIC STATE WITH ACUTE EXACERBATION |
| 295.35 | PARANOID TYPE SCHIZOPHRENIA, IN REMISSION |
| 295.4 | ACUTE SCHIZOPHRENIC EPISODE |
| 295.40 | ACUTE SCHIZOPHRENIC EPISODE, UNSPECIFIED STATE |
| 295.41 | ACUTE SCHIZOPHRENIC EPISODE, SUBCHRONIC STATE |
| 295.42 | ACUTE SCHIZOPHRENIC EPISODE, CHRONIC STATE |
| 295.43 | ACUTE SCHIZOPHRENIC EPISODE, SUBCHRONIC STATE WITH ACUTE EXACERBATION |
| 295.44 | ACUTE SCHIZOPHRENIC EPISODE, CHRONIC STATE WITH ACUTE EXACERBATION |
| 295.45 | ACUTE SCHIZOPHRENIC EPISODE, IN REMISSION |
| 295.5 | SCHIZOTYPAL DISORDER-LATENT SCHIZOPHRENIA |
| 295.50 | SCHIZOTYPAL DISORDER-LATENT SCHIZOPHRENIA, UNSPECIFIED STATE |
| 295.52 | SCHIZOTYPAL DISORDER-LATENT SCHIZOPHRENIA, CHRONIC STATE |
| 295.53 | SCHIZOTYPAL DISORDER-LATENT SCHIZOPHRENIA, SUBCHRONIC STATE WITH ACUTE EXACERBATION |
| 295.54 | SCHIZOTYPAL DISODER-LATENT SCHIZOPHRENIA, CHRONIC STATE WITH ACUTE EXACERBATION |
| 295.55 | SCHIZOTYPAL DISORDER-LATENT SCHIZOPHRENIA, IN REMISSION |
| 295.6 | RESIDUAL SCHIZOPHRENIA |
| 295.60 | RESIDUAL SCHIZOPHRENIA, UNSPECIFIED STATE |
| 295.62 | RESIDUAL SCHIZOPHRENIA, CHRONIC STATE |
| 295.64 | RESIDUAL SCHIZOPHRENIA, CHRONIC STATE WITH ACUTE EXACERBATION |
| 295.65 | RESIDUAL SCHIZOPHRENIA, IN REMISSION |
| 295.7 | SCHIZO-AFFECTIVE TYPE SCHIZOPHRENIA |
| 295.70 | SCHIZO-AFFECTIVE TYPE SCHIZOPHRENIA, UNSPECIFIED STATE |
| 295.71 | SCHIZO-AFFECTIVE TYPE SCHIZOPHRENIA, SUBCHRONIC STATE |
| 295.72 | SCHIZO-AFFECTIVE TYPE SCHIZOPHRENIA, CHRONIC STATE |
| 295.73 | SCHIZO-AFFECTIVE TYPE SCHIZOPHRENIA, SUBCHRONIC STATE WITH ACUTE EXACERBATION |
| 295.74 | SCHIZO-AFFECTIVE TYPE SCHIZOPHRENIA, CHRONIC STATE WITH ACUTE EXACERBATION |
| 295.75 | SCHIZO-AFFECTIVE TYPE SCHIZOPHRENIA, IN REMISSION |
| 295.8 | OTHER SPECIFIED TYPES OF SCHIZOPHRENIA |
| 295.80 | OTHER SPECIFIED TYPES OF SCHIZOPHRENIA, UNSPECIFIED STATE |
| 295.82 | OTHER SPECIFIED TYPES OF SCHIZOPHRENIA, CHRONIC STATE |
| 295.83 | OTHER SPECIFIED TYPES OF SCHIZOPHRENIA, SUBCHRONIC STATE WITH ACUTE EXACERBATION |
| 295.84 | OTHER SPECIFIED TYPES OF SCHIZOPHRENIA, CHRONIC STATE WITH ACUTE EXACERBATION |
| 295.85 | OTHER SPECIFIED TYPES OF SCHIZOPHRENIA, IN REMISSION |
| 295.9 | UNSPECIFIED SCHIZOPHRENIA |
| 295.90 | UNSPECIFIED TYPE SCHIZOPHRENIA, UNSPECIFIED STATE |
| 295.91 | UNSPECIFIED TYPE SCHIZOPHRENIA, SUBCHRONIC STATE |
| 295.92 | UNSPECIFIED TYPE SCHIZOPHRENIA, CHRONIC STATE |
| 295.920 | OBSESSIVE-COMPULSIVE DISORDER, UNSPECIFIED |
| 295.93 | UNSPECIFIED TYPE SCHIZOPHRENIA, SUBCHRONIC STATE WITH ACUTE EXACERBATION |
| 295.94 | UNSPECIFIED TYPE SCHIZOPHRENIA, CHRONIC STATE WITH ACUTE EXACERBATION |
| 332.1 | PARKINSONISM, SECONDARY |
| 332.10 | PARKINSONISM DUE TO DRUGS |
| 332.11 | TARDIVE DYSKINESIA |
| 333.99 | RESTLESS LEGS |
| 333.990 | PERIODIC LEGS MOVEMENTS |
| 431 | INTRACEREBRAL HEMORRHAGE |
| 434.01 | CEREBRAL THROMBOSIS WITH CEREBRAL INFARCTION |
| 434.11 | CEREBRAL EMBOLISM WITH CEREBRAL INFARCTION |
| 434.9 | CEREBRAL ARTERY OCCLUSION, UNSPECIFIED |
| 434.90 | ISCHEMIC CEREBROVASCULAR DISEASE (CVA) WITHOUT MENTION OF INFARCTION |
| 434.91 | ISCHEMIC CEREBROVASCULAR DISEASE (CVA) WITH INFARCTION |
| 436 | ACUTE, BUT ILL-DEFINED, CEREBROVASCULAR DISEASE-CVA |
| 436.3 | STROKE |
| 431 | INTRACEREBRAL HEMORRHAGE |
| 434.01 | CEREBRAL THROMBOSIS WITH CEREBRAL INFARCTION |
| 434.11 | CEREBRAL EMBOLISM WITH CEREBRAL INFARCTION |
| 434.9 | CEREBRAL ARTERY OCCLUSION, UNSPECIFIED |
| 434.90 | ISCHEMIC CEREBROVASCULAR DISEASE (CVA) WITHOUT MENTION OF INFARCTION |
| 434.91 | ISCHEMIC CEREBROVASCULAR DISEASE (CVA) WITH INFARCTION |
| 436 | ACUTE, BUT ILL-DEFINED, CEREBROVASCULAR DISEASE-CVA |
| 436.3 | STROKE |

Table S9.

List of antipsychotic medications which purchase prior to the index date prevents inclusion in the cohort

| N05AA01 | Chlorpromazine |
| --- | --- |
| N05AA02 | Levomepromazine |
| N05AB02 | Fluphenazine |
| N05AB03 | Perphenazine |
| N05AB06 | Trifluoperazine |
| N05AB08 | Thioproperazine |
| N05AC01 | Periciazine |
| N05AC02 | Thioridazine |
| N05AD01 | Haloperidol |
| N05AE03 | Sertindole |
| N05AE04 | Ziprasidone |
| N05AF01 | Flupentixol |
| N05AF03 | Chlorprothixene |
| N05AF05 | Zuclopenthixol |
| N05AG02 | Pimozide |
| N05AG03 | Penfluridol |
| N05AH02 | Clozapine |
| N05AH03 | Olanzapine |
| N05AH04 | Quetiapine |
| N05AH05 | Asenapine |
| N05AH06 | Clotiapine |
| N05AL01 | Sulpiride |
| N05AL03 | Tiapride |
| N05AL05 | Amisulpride |
| N05AN01 | Lithium |
| N05AX08 | Risperidone |
| N05AX12 | Aripiprazole |
| N05AX13 | Paliperidone |
| N05AX14 | Iloperidone |
| N05AX15 | Cariprazine |
| N05AX16 | Brexpiprazole |

Table S10.

List of variables included in the disease severity model. For each medication we compute the number of units purchased each year the annual score.

| **Gender of the patient (1 for female)** |  |  |  |  |
| --- | --- | --- | --- | --- |
| Medication (yearly quantity of medication unit for each of the following catalog entries) | ATC code | ATC description | Serving Form | Giving Form |
| AMANDIN 100 MG 100 TAB | N04BB01 | Amantadine | TB | PO |
| AMANTADIN HX 100MG 30 29G | N04BB01 | Amantadine | TB | PO |
| AMANTADINE 100MG 100T 29G | N04BB01 | Amantadine | TB | PO |
| AMANTADINSULFAT 200MG_500 | N04BB01 | Amantadine | IJ | IV |
| APO-GO 10 MG/ML 5ML 5 AMP | N04BC07 | Apomorphine | IJ | SC |
| APO-GO 20 MG/2ML 5 AMP | N04BC07 | Apomorphine | IJ | SC |
| APO-GO PEN 10 MG_ML 3ML*5 | N04BC07 | Apomorphine | SR | SC |
| AZILECT 1 MG 30 TAB | N04BD02 | Rasagiline | TB | PO |
| COMTAN 200 MG 30 TAB | N04BX02 | Entacapone | TB | PO |
| DEKINET 2 MG 30 TAB | N04AA02 | Biperiden | TB | PO |
| DOPICAR 30 TAB | N04BA02 | Levodopa and Decarbo | TB | PO |
| DUODOPA GEL 100 ML 7 CASS | N04BA02 | Levodopa and Decarbo | GE | IS |
| ENTACAPONE 200 MG 30 TAB | N04BX02 | Entacapone | TB | PO |
| JUMEX 5 50 TAB | N04BD01 | Selegiline | TB | PO |
| KEMADRIN 5 MG 100 TAB | N04AA04 | Procyclidine | TB | PO |
| LEVOPAR PLUS 125 MG 30 C | N04BA02 | Levodopa and Decarbo | TB | PO |
| LEVOPAR PLUS 250 MG 30 C | N04BA02 | Levodopa and Decarbo | TB | PO |
| MADOPAR 100/25MG CAP 29-G | N04BA02 | Levodopa and Decarbo | CP | PO |
| MADOPAR 200/50MG C MBI 29 | N04BA02 | Levodopa and Decarbo | CP | PO |
| MADOPAR 200/50MG CAP 29-G | N04BA02 | Levodopa and Decarbo | CP | PO |
| PARILAC 10 MG 20 CAP | N04BC01 | Bromocriptine | CP | PO |
| PARITREL 20 TAB | N04BB01 | Amantadine | TB | PO |
| PARTANE 2 MG 50 TAB | N04AA01 | Trihexyphenidyl | TB | PO |
| PARTANE 5 MG 50 TAB | N04AA01 | Trihexyphenidyl | TB | PO |
| PERGOLIDE 0.05 MG 100 TA | N04BC02 | Pergolide | TB | PO |
| PERGOLIDE 0.25 MG 100 T | N04BC02 | Pergolide | TB | PO |
| PERGOLIDE TEVA 0.25MG 100 | N04BC02 | Pergolide | TB | PO |
| PK MERZ 100 MG TAB 29-G | N04BB01 | Amantadine | TB | PO |
| PK MERZ 100 TAB 29-G | N04BB01 | Amantadine | TB | PO |
| PK MERZ 100MG TAB RAZ 29G | N04BB01 | Amantadine | TB | PO |
| PK MERZ INF 200MG/500ML*2 | N04BB01 | Amantadine | IJ | IV |
| PRAMIPEXOLE TEVA 0.25 MG | N04BC05 | Pramipexole | TB | PO |
| PRAMIPEXOLE TEVA 1MG 100T | N04BC05 | Pramipexole | TB | PO |
| RASAGILINE-TRIMA 1 MG 30 | N04BD02 | Rasagiline | TB | PO |
| REQUIP 0.25 MG 21 TAB | N04BC04 | Ropinirole | TB | PO |
| REQUIP 0.25 MG 210 TAB | N04BC04 | Ropinirole | TB | PO |
| REQUIP 0.5 MG 21 TAB | N04BC04 | Ropinirole | TB | PO |
| REQUIP 1 MG 21 TAB | N04BC04 | Ropinirole | TB | PO |
| REQUIP 1 MG 84 TAB | N04BC04 | Ropinirole | TB | PO |
| REQUIP 2 MG 21 TAB | N04BC04 | Ropinirole | TB | PO |
| REQUIP 2 MG 84 TAB | N04BC04 | Ropinirole | TB | PO |
| REQUIP 5 MG 21 TAB | N04BC04 | Ropinirole | TB | PO |
| REQUIP 5 MG 84 TAB | N04BC04 | Ropinirole | TB | PO |
| REQUIP MODUTAB 2 MG 28TAB | N04BC04 | Ropinirole | TB | PO |
| REQUIP MODUTAB 4 MG 28TAB | N04BC04 | Ropinirole | TB | PO |
| REQUIP MODUTAB 8 MG 28TAB | N04BC04 | Ropinirole | TB | PO |
| RODENAL 2 MG 30 TAB | N04AA01 | Trihexyphenidyl | TB | PO |
| RODENAL 5 MG 30 TAB | N04AA01 | Trihexyphenidyl | TB | PO |
| ROPINIROLE 0.25 MG 210 T | N04BC04 | Ropinirole | TB | PO |
| ROPINIROLE 0.5 MG 21 TAB | N04BC04 | Ropinirole | TB | PO |
| ROPINIROLE 1 MG 84 TAB | N04BC04 | Ropinirole | TB | PO |
| ROPINIROLE 2 MG 84 TAB | N04BC04 | Ropinirole | TB | PO |
| ROPINIROLE 5 MG 84 TAB | N04BC04 | Ropinirole | TB | PO |
| SELEGILINA GENER 5MG 29-G | N04BD01 | Selegiline | TB | PO |
| SELEGILINE 5 MG 50 T CTS | N04BD01 | Selegiline | TB | PO |
| SIFROL 0.25 MG 100 TAB | N04BC05 | Pramipexole | TB | PO |
| SIFROL 1.0 MG 100 TAB | N04BC05 | Pramipexole | TB | PO |
| SIFROL ER 0.375 MG 30 TAB | N04BC05 | Pramipexole | TB | PO |
| SIFROL ER 0.75 MG 30 TAB | N04BC05 | Pramipexole | TB | PO |
| SIFROL ER 1.5 MG 30 TAB | N04BC05 | Pramipexole | TB | PO |
| SINEMET CR 30 TAB | N04BA02 | Levodopa and Decarbo | TB | PO |
| SINEMET CR 50_200 100 TAB | N04BA02 | Levodopa and Decarbo | TB | PO |
| SINEMET CR 50_200 60T 29G | N04BA02 | Levodopa and Decarbo | TB | PO |
| SINEMET RETARD 50/200 100 | N04BA02 | Levodopa and Decarbo | TB | PO |
| STALEVO 100/25/200 MG 30T | N04BA03 | Levodopa, Decarboxyl | TB | PO |
| STALEVO 125/31.25/200 30T | N04BA03 | Levodopa, Decarboxyl | TB | PO |
| STALEVO 150/37.5/200MG 30 | N04BA03 | Levodopa, Decarboxyl | TB | PO |
| STALEVO 200/50/200 MG 30T | N04BA03 | Levodopa, Decarboxyl | TB | PO |
| STALEVO 50/12.5/200 MG 30 | N04BA03 | Levodopa, Decarboxyl | TB | PO |
| STALEVO 75/18.75/200MG 30 | N04BA03 | Levodopa, Decarboxyl | TB | PO |
| TRIMEXOL 0.25 MG 100 TAB | N04BC05 | Pramipexole | TB | PO |
| TRIMEXOL 1 MG 100 TAB | N04BC05 | Pramipexole | TB | PO |
| XADAGO 100 MG 30 TAB | N04BD03 | Safinamide | TB | PO |
| XADAGO 50 MG 30 TAB | N04BD03 | Safinamide | TB | PO |
